# Genetic Architecture of Circulating Metabolic Biomarkers Across Ancestral Populations

**DOI:** 10.64898/2025.12.03.25341540

**Authors:** Yohei Takeuchi, Masaru Koido, Yunye He, Shota Nakamura, Xin Liu, Naoki Itokawa, Hiromi Tsuru, Yoji Sagiya, Akiko Nagai, Takayuki Morisaki, Koichi Matsuda, Yoichiro Kamatani

## Abstract

Circulating metabolite levels hold promise for disease risk prediction, yet the genetic architecture and cross-ancestry transferability remain incompletely characterized. Here, we conducted the largest genome-wide association study in East Asians to date, quantifying 248 blood metabolic biomarkers using nuclear magnetic resonance spectroscopy in 37,969 Japanese individuals. We identified eight novel associations and pinpointed rs75326924, causing CD36 deficiency, as a putative causal variant influencing average diameter for low-density lipoprotein particles. A cross-ancestry meta-analysis of 657,341 individuals uncovered 28 additional novel associations and 11,167 credible sets, indicating 52.6% of ancestry-specific causal variants despite highly correlated marginal effect sizes (Pearson r = 0.78 ‒ 0.98). Integration with liver single-nucleus transcriptomics highlighted perivascular hepatocytes as a key cell type of the genetic regulation of circulating biomarkers. Finally, weighted sums of metabolic biomarkers derived from the UK Biobank predicted myocardial infarction and cirrhosis risks in Japanese individuals. Our findings refine the genetic architecture of metabolic biomarkers and underscore their clinical relevance across ancestries.

## Main

Metabolites are low molecular weight compounds. In organisms, they are synthesized and degraded in cellular and tissue-level metabolic pathways, and perturbations in these processes can influence disease risk. Metabolite levels in tissues and biofluids capture these dynamics^1^, and blood metabolic biomarker profiling, in particular, provides a snapshot of an individual’s metabolic state. Genetic factors account for approximately 20–80% of the variation in metabolic levels^2^. Thus, elucidating the genetic architecture of metabolites is essential for uncovering the biological underpinnings of human metabolism.

Large-scale metabolomic genome-wide association studies (GWASs) (N > 10,000) have been conducted in several European cohorts^3–5^, which have investigated the genetic factors and elucidated new metabolic processes. On the other hand, because genetic association signals strongly depend on linkage disequilibrium (LD) structure and allele frequencies, GWASs in non-European populations have the potential to identify novel loci undetected in European studies. Despite this advantage, large-scale metabolomic GWASs in the East Asian population have been limited to a study in Japan^6^ and two studies in China^7,8^, each involving 10,333, 10,792, and 34,394 individuals, respectively. Therefore, studies with larger East Asian sample sizes are necessary to comprehensively elucidate the metabolomic genetic architecture.

Previous studies have shown that measurements of metabolic biomarkers by nuclear magnetic resonance (NMR) spectroscopy and their weighted sums were associated with current and future health status^9–12^. Among the metabolomics platforms, NMR spectroscopy generally exhibits fewer batch effects and lower costs than other methods, such as mass spectrometry (MS)^13^, making NMR-derived biomarker panels attractive for clinical applications. However, environmental and genetic determinants of disease risk, as well as disease prevalence and primary etiologies, differ across ancestral populations. Consequently, caution is required when interpreting the predictive ability for disease risk across different ancestries. For example, a model trained in a European population demonstrated poor lung cancer predictive accuracy among Asian ever-smokers^14^. Accordingly, the relationship between metabolic biomarkers and disease risk may also vary between European and East-Asian populations, and the findings derived from European studies cannot be directly generalized to East-Asian populations^11^.

In this study, we performed a GWAS of 248 metabolic biomarkers, including amino acids, lipids, and lipoproteins, analyzed in blood samples from 37,969 Japanese participants in Biobank Japan (BBJ) using NMR spectroscopy. This study is the largest metabolomic GWAS in the East Asian population to date. Using the metabolomic GWAS results from BBJ and UK Biobank (UKB) and Estonian Biobank (EBB), encompassing a combined N = 657,341 individuals, we characterized the ancestry-specific and shared genetic effects on metabolic biomarkers. To map these genetic signals to biology, we linked the metabolomic GWAS results to transcriptome profiles from a range of tissues and cell populations most significantly associated with each biomarker. Finally, utilizing metabolomic measurements and clinical data from BBJ, we evaluated the prognostic value of metabolic biomarkers for disease risk in the Japanese population.

## Results

A total of 250 metabolic biomarkers (Supplementary Table 1) were initially analyzed using the NMR platform in 45,270 serum samples from the BBJ first cohort (BBJ1), which consisted of two independently genotyped sub-cohorts: BBJ1-180K, with 182,536 individuals, and BBJ1-12K, with 11,716 individuals. Glycerol and 3-hydroxybutyrate were excluded from this study because available measurements were less than 90% of the total samples. 248 analyzed metabolic biomarkers were classified into 18 groups based on their biological functions. For the present study, we used samples that passed both metabolomic and genotype quality control (QC) (see Methods). After QC, 37,969 samples from BBJ1-180K were used for metabolomic GWAS, 3,522 samples from BBJ1-12K were used for additional genetic association tests, and 15,137 to 34,765 samples from BBJ1-180K were available for survival analysis evaluating the associations between metabolic biomarker(s) and disease risk.

### Investigation of genetic effects on metabolic biomarkers in the Japanese population

Using 37,969 Japanese participants from BBJ1-180K, we investigated the associations between 11 million imputed variants (minor allele frequency (MAF) > 0.1 % and an imputation quality score *R*^2^ > 0.7) and 248 metabolic biomarkers using a generalized linear mixed model implemented in BOLT-LMM^15^. To account for multiple testing across 248 metabolic biomarkers, we applied a study-wide significance threshold (*P* value < 5 × 10^−8^/20, Bonferroni correction; see Methods) in the metabolomic GWAS. We identified 2,140 associations involving 297 unique lead variants and 244 metabolic biomarkers and detected 76 regions (one to 34 unique lead variants per region) without overlap across metabolic biomarkers (Fig. 1a and Supplementary Table 2). Of these, eight pairs (involving six unique lead variants and eight metabolic biomarkers) were novel, corresponding to six loci across metabolic biomarkers (Table 1).

**Fig. 1.**
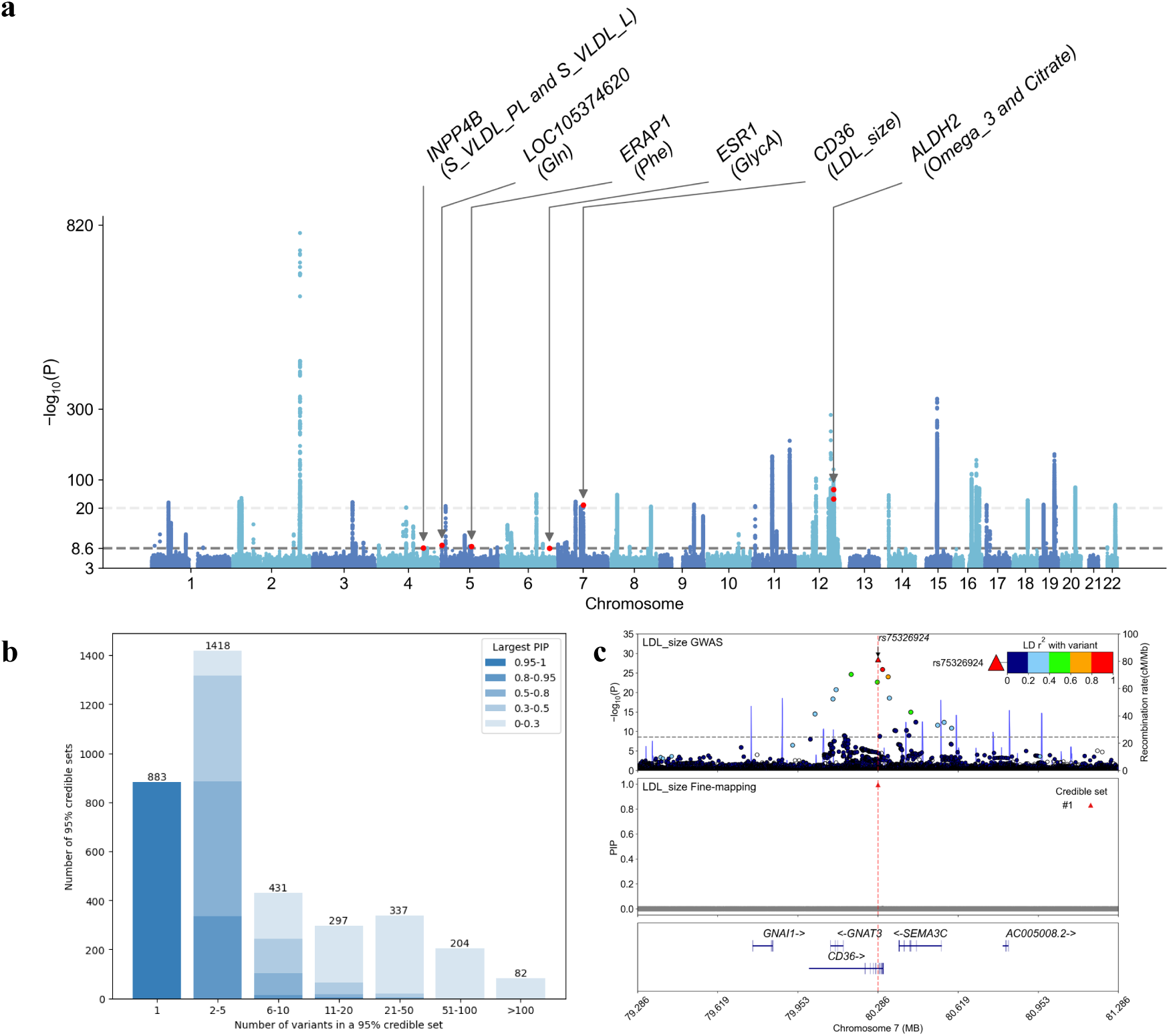
GWAS results in BBJ1-180K. **a**, Manhattan plot of the metabolomic GWAS in BBJ1-180K. Association results for 248 metabolic biomarkers are over-laid into a single panel. For variants above the blue dashed line, values are rescaled. The gray dashed line denotes the study-wide significance threshold (*P* value < 5 × 10^−8^/20). Red dots mark variants that represent novel associations; each dot is annotated with the region name and corresponding metabolic biomarker. Abbreviations of metabolic biomarkers are listed in Supplementary Table 1. **b**, Number of variants and the maximum PIP within 3,652 95% credible sets identified by fine-mapping. Each bin is colored according to the number the largest PIP in the credible set (The larger PIP, blue is darker.). **c**, Regional plot of the *CD36* locus for LDL size from GWAS (top) and fine-mapping (bottom) results in BBJ1-180K. In the top panel, the dotted line shows the study-wide significance threshold (*P* value < 5 × 10^−8^/20). LD with rs75326924, calculated based on 1KGP3 EAS, is shown by color scale.

**Table 1.** Eight novel associations identified by GWAS in BBJ1-180K.

| Metabolic | rs ID | Chromosome | Position (GRCh37) | Region name | Effect allele/ noneffect allele | Beta | SE | P value | EAF | EAF_1KG_EUR |
| --- | --- | --- | --- | --- | --- | --- | --- | --- | --- | --- |
| Total lipids in small VLDL | rs1497401 | 4 | 143321490 | *INPP4B* | T/C | 0.056 | 0.0092 | 2.00E-09 | 0.302 | 0.368 |
| Phospholipids in small VLDL | rs1497401 | 4 | 143321490 | *INPP4B* | T/C | 0.056 | 0.0091 | 1.80E-09 | 0.302 | 0.368 |
| Glutamine | rs56981040 | 5 | 2992414 | *LOC105374620* | G/T | -0.061 | 0.0096 | 3.10E-10 | 0.828 | 0.974 |
| Phenylalanine | rs70981840 | 5 | 96119579 | *ERAP1* | A/ACT | 0.044 | 0.0073 | 6.90E-10 | 0.378 | 0.147 |
| Glycoprotein acetyls | rs560647736 | 6 | 152278333 | *ESR1* | C/T | 0.64 | 0.11 | 2.30E-09 | 0.999 | 1 |
| Average diameter for LDL particles | rs75326924 | 7 | 80286003 | *CD36* | C/T | 0.23 | 0.020 | 3.20E-29 | 0.955 | 1 |
| Omega-3 fatty acids | rs149212747 | 12 | 111836771 | *ALDH2* | A/AC | 0.16 | 0.0088 | 2.20E-73 | 0.754 | 1 |
| Citrate | rs149212747 | 12 | 111836771 | *ALDH2* | A/AC | -0.13 | 0.0088 | 1.10E-46 | 0.754 | 1 |

SE: Standard error. EAF: Effect allele frequency. EAF\_BBJ: EAF in BBJ1-180K. EAF\_EUR: EAF in Europeans of 1KGP3.

To replicate these eight novel associations, we analyzed BBJ1-12K (N = 3,528). Six associations showed concordant effect directions with the discovery GWAS results, although the sign test did not reach nominal significance (*P* value = 0.14). Furthermore, three associations (the *CD36* region with the average diameter for low-density lipoprotein particles (LDL size) and the *ALDH2* region with omega-3 fatty acids and citrate) showed nominal significance (*P* value < 0.05) in the same direction, with three of these remaining significant after Bonferroni correction (*P* value < 0.05/8) (Supplementary Table 3).

To find putative causal variants for metabolic biomarkers, we performed statistical fine-mapping of 2 Mb regions centered on each lead variant or index variant from secondary signals (see Methods). We merged partially overlapping regions corresponding to the same metabolic biomarker, resulting in 2,111 regions comprising 285 lead or index variants and 244 metabolic biomarkers. Using SuSiE-RSS^16^, we identified 3,652 independent causal signals (95% credible sets; CSs) across 244 metabolic biomarkers (Fig. 1b). Each CS contained one to 203 variants (median: 4). 883 CSs (239 metabolic biomarkers and 61 variants) comprised a single variant with a posterior inclusion probability (PIP) > 0.95, and 1,240 CSs (242 metabolic biomarkers and 89 variants) harbored at least one variant with PIP > 80%. Among the 89 variants, 24 directly affect the protein function (functional variants; see Methods), indicating that the proteins encoded by the corresponding 20 genes alter metabolite levels (Supplementary Table 4). Notably, these coding variants included rs75326924, a missense variant in *CD36* (Fig. 1c). This variant is known to cause CD36 deficiency and is also associated with atherosclerotic cardiovascular disease^17^. rs75326924 was fine-mapped as a causal variant for 15 metabolic biomarkers, including a novel association with LDL size (PIP = 1.0). For the remaining 65 fine-mapped variants, we evaluated whether they were located within the enhancer elements linking to target genes in 131 human tissue- or cell-types predicted by the Activity-by-Contact (ABC) model^18,19^ (ABC score > 0.015). This analysis identified 23 variants in 155 elements targeting 65 genes in 106 tissue or cell-types (Supplementary Table 5). Among non-coding causal variants, we identified rs77869005, 5’ untranslated region in *HAL*, as a potential causal variant for histidine levels (PIP = 0.99). The MAF of this variant was markedly higher in the East Asian population, particularly in the Japanese population, than in the European population (MAF in 1000 genomes project phase3 (1KGP3)^20^ Europeans (EUR) = 0.0010, 1KGP3 East Asians (EAS) = 0.051; 1KGP3 Japanese in Tokyo = 0.082, and BBJ1-180K = 0.084), suggesting that its higher frequency in the Japanese population enabled the finding in our study. The ABC model predicts that elements containing rs77869005 influence expression levels of *HAL* in 12 cell and tissue types, with the highest ABC score observed in the liver. Histidine ammonia-lyase coded by *HAL* is the enzyme that converts histidine to urocanic acid in the liver^21^. These results suggested that rs77869005 influences blood histidine levels by regulating the *HAL* enhancer activity in the liver.

### Meta-analysis and comparisons of genetic effect sizes across ancestral populations

To investigate genetic effects on metabolic biomarkers shared across ancestral populations, we performed a meta-analysis of 8 million imputed variants and 248 metabolic biomarkers using a fixed effects model with the inverse variance weighted (IVW) method implemented in METAL^22^. We integrated the metabolomic GWAS results in BBJ1-180K with the cross-ancestry meta-analysis results from UKB and EBB (UKB-EBB)^23^ (N_total_ = 657,272). After applying the study-wide Bonferroni correction (*P* value < 5 × 10^−8^/20), we identified 44,976 associations (7,983 unique lead variants and 248 metabolic biomarkers) and 665 regions with no overlap across metabolic biomarkers (Supplementary Table 6). Of these, 28 associations (16 unique lead variants and 26 metabolic biomarkers) and 14 regions across metabolic biomarkers were identified as novel associations in addition to those detected by GWAS in BBJ1-180K (Table 2).

**Table 2.**
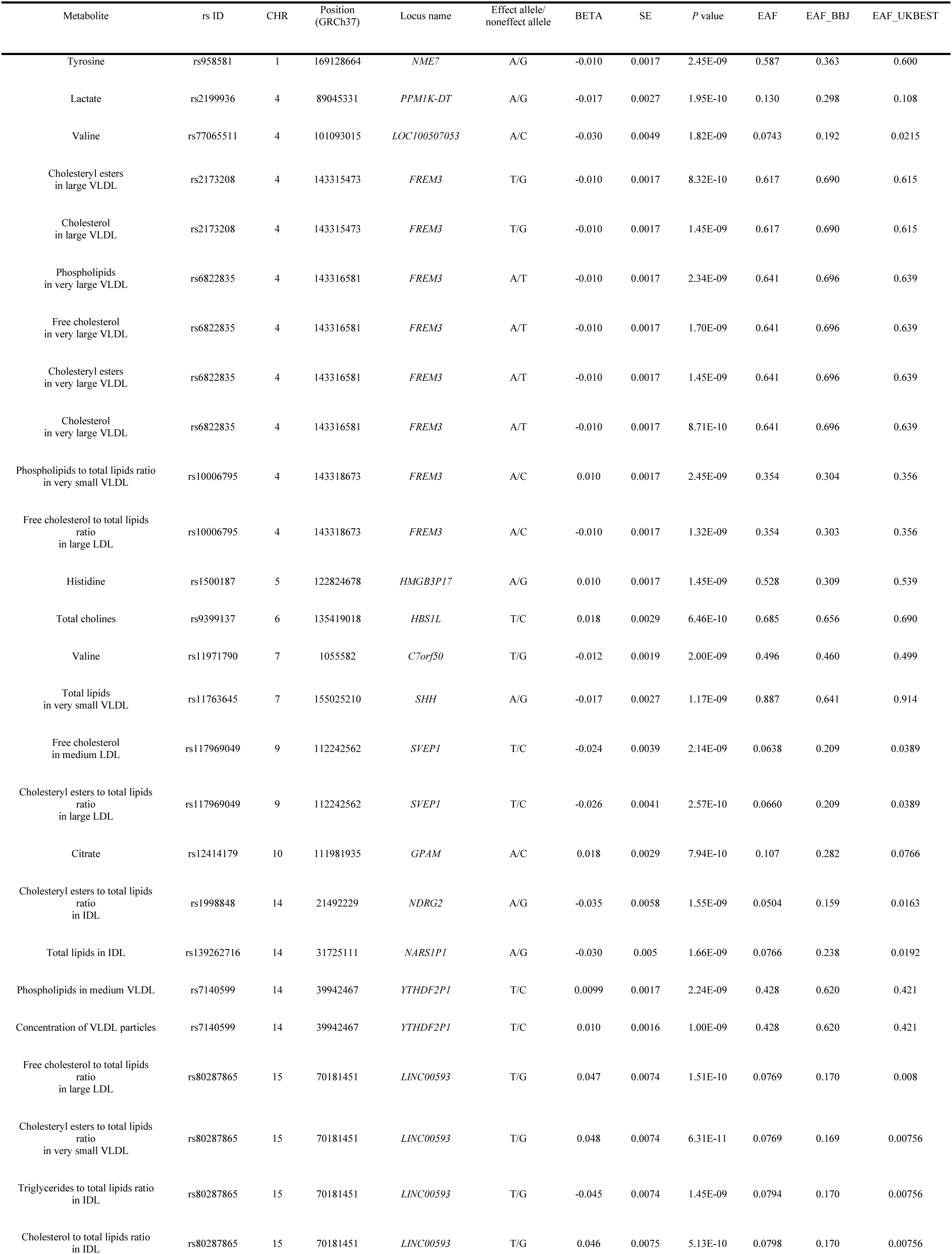
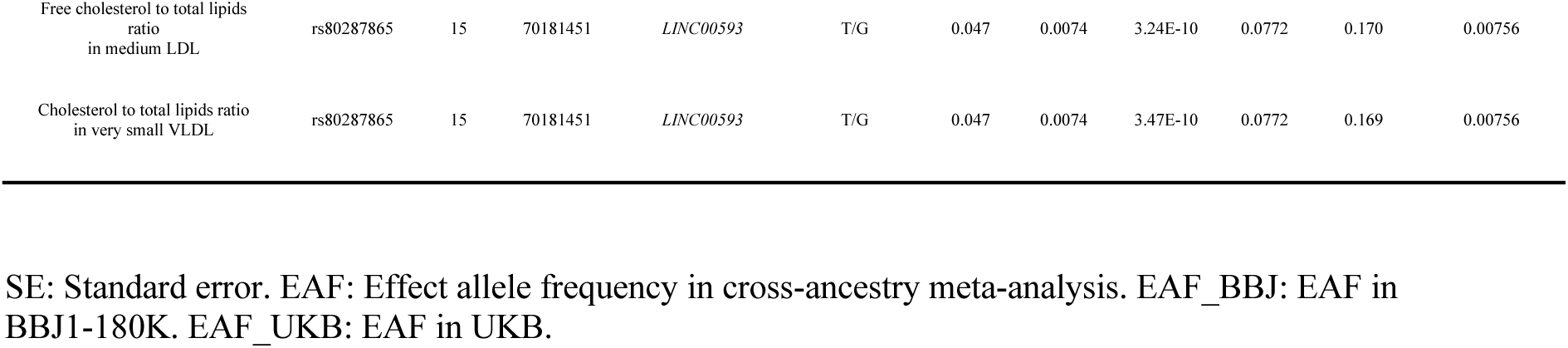
28 novel associations identified by cross-ancestry meta-analysis.

To replicate these 28 novel associations, we evaluated the corresponding associations in BBJ1-12K. 19 associations showed concordant effect directions with the discovery meta-analysis results, and a sign test revealed a statistically significant enrichment of consistent effects (*P* value = 0.044) (Supplementary Table 7). Since statistical powers in BBJ1-12K were low (Power = 0.064-0.27) (Supplementary Table 7), we further evaluated these novel associations using almost independent large-scale cross-ancestry meta-analysis results (N_total_ = 136,016), with partial overlap with our meta-analysis (N = 3,701 in EBB)^3^. Of the 28 novel associations, 27 showed consistent effect directions in the large-scale meta-analysis, and a sign test again confirmed significant enrichment (*P* value = 1.1 × 10^−7^) (Supplementary Table 7). In addition, 11 associations showed nominal significance (*P* value < 0.05) in the same direction; one of these remained significant after Bonferroni correction (*P* value < 0.05/28, Supplementary Table 7).

We assessed the cross-population similarity of genetic effects on metabolic biomarkers by evaluating their correlations between the East Asian population from BBJ1-180K and six ancestral populations from UKB-EBB or UKB: the European population from UKB-EBB, and the Central/South Asian, African, East Asian, Middle Eastern, and Admixed American populations from UKB. First, we extracted the lead variants identified in BBJ1-180K. We compared their effect sizes with those in each UKB-EBB or UKB population, using variants with MAF > 1% in both populations being compared. The Pearson correlation coefficients of effect sizes ranged from 0.78 in Admixed Americans to 0.95 in Europeans, with a relatively high correlation of 0.93 observed in UKB East Asians (Fig. 2a and Supplementary Table 8). Next, we performed a reverse comparison: for each UKB-EBB or UKB population, we extracted lead variants associated with metabolic biomarkers. We compared their effect sizes with those in BBJ1-180K, again using variants with MAF > 1% in both populations. Pearson correlation coefficients ranged from 0.85 in Middle Easterns to 0.98 in UKB East Asians (Fig. 2b and Supplementary Table 9). These results indicate that the genetic effects on metabolic biomarkers are largely consistent across populations, particularly between East Asians in BBJ1-180K and those in UKB.

**Fig. 2.**
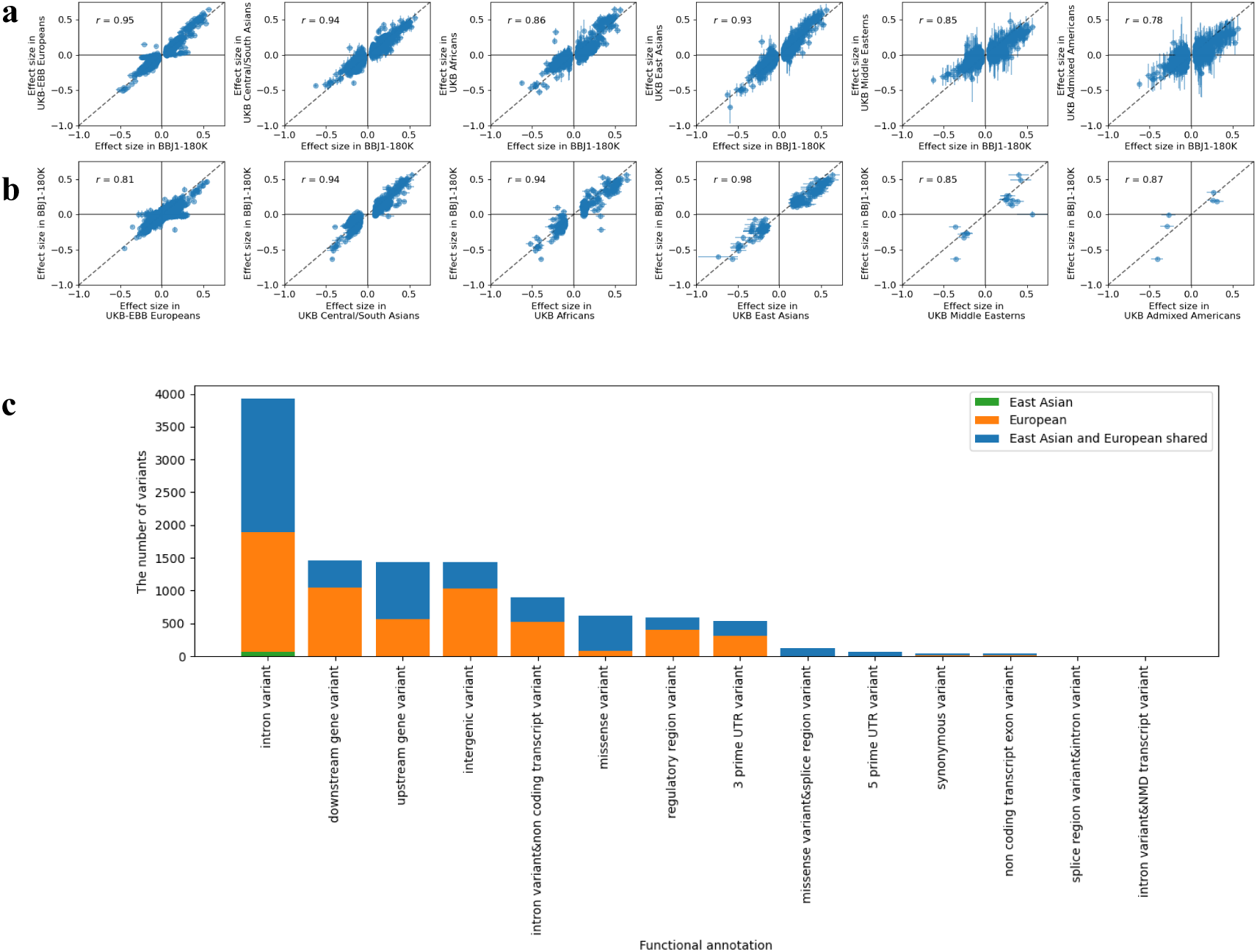
Comparison of genetic effects on metabolic biomarkers across ancestral populations. **a**, **b** Effect size correlations for the lead variants identified in (**a**) BBJ1-180K or (**b**) UKB-EBB or UKB. For each panel the effect sizes in BBJ1-180K are plotted against those in the six ancestral populations of UKB. *r*: Pearson’s correlation. **c,** Stacked bar plot of 11,167 putative causal variants in either population or both populations across 14 types of functional annotations.

Next, we performed a multi-ancestry fine-mapping using MESuSiE^24^, integrating the GWAS results from BBJ1-180K and the European-specific meta-analysis from UKB-EBB. We focused on 2,111 regions associated with traits in BBJ1-180K and identified 14,266 95% CSs across 244 metabolic biomarkers. Each of the 11,167 CSs contained at least one variant with a PIP > 0.5 for being ancestry-specific or shared (Supplementary Table 10). Despite the largely consistent effect sizes of lead variants between the East Asian and European populations, we estimated 5,881 variants (52.6%) to be population-specific causal variants, comprising 5,802 European-specific and 79 East Asian-specific causal variants. The 3,099 remaining CSs did not contain a variant with PIP > 0.5 for being ancestry-specific or shared. To investigate the potential functional mechanisms, we examined variants with the highest PIP ( > 0.5) in 95% CSs and found that population-shared variants were more prevalent in categories including missense variants among the 14 types of annotation, while the most frequently observed variants were intronic (Fig. 2c and Supplementary Table 11).

### Genetic links to blood metabolic biomarkers and tissue- and cell-types

To identify tissue-type specificity of genetic associations for metabolic biomarkers, we investigated the gene-level associations between circulating 248 metabolic biomarkers and 30 types of tissues using data from the Genotype-Tissue Expression (GTEx)^25^ using MAMGA^26^ implemented in FUMA (v.1.6.2)^27^. Using the GWAS results from BBJ1-180K, we identified five significant biomarker-tissue pairs, all of which were for the liver (*P* value < 0.05/(20×30) after Bonferroni correction; see Methods). Using the European-specific meta-analysis results from UKB-EBB, we identified 252 significant pairs (227 metabolic biomarkers and seven tissues), including the same five pairs identified in BBJ1-180K (*P* value < 0.05/(20×30), after Bonferroni correction; see Methods) (Fig. 3a and Supplementary Table 12). Across all analyzed tissues, the liver showed the largest number of associations (222 metabolic biomarkers). Z-scores for these liver-biomarker associations were significantly higher than those for the other 29 tissues (t-test *P* value < 0.05/(29 × 2), after Bonferroni correction; Supplementary Fig. 1) in both BBJ1-180K and UKB-EBB, suggesting the central role of the liver in the genetic regulation of these metabolic biomarkers in both populations. Beyond the liver, the small intestine was associated with 13 metabolic biomarkers, whole blood with 10, the spleen with three, the skeletal muscle with two, and the kidney and thyroid with creatinine.

**Fig. 3.**
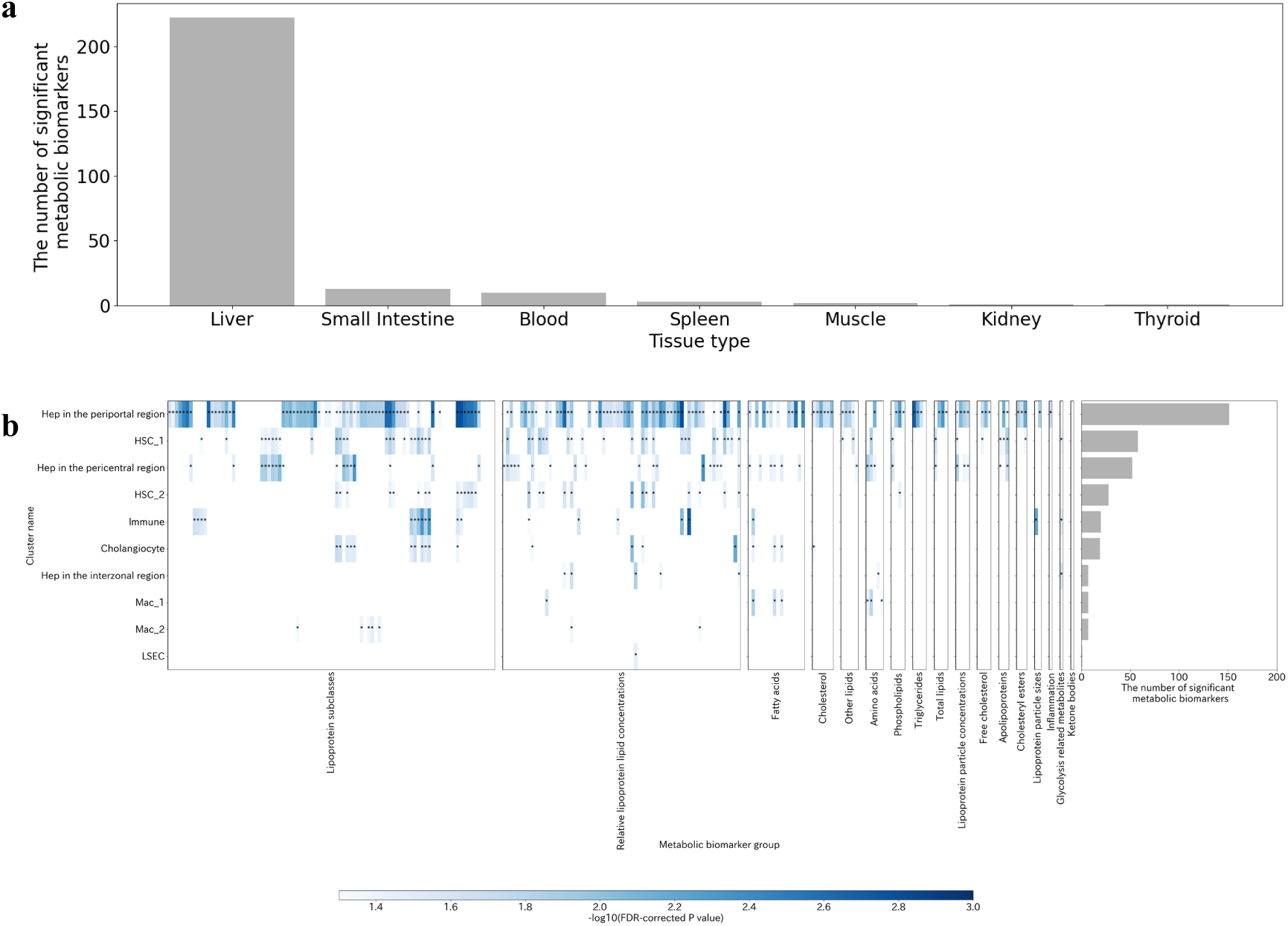
Gene-level association landscape for metabolic biomarkers across tissues and cell types in the liver. **a**, Bar plot of the number of significant metabolic biomarkers (P value < 0.05 / (20×30) ) for each tissue. Only the seven tissues out of the 30 whole-body tissues examined that yielded at least one significant biomarker are shown. **b**, Composite view of the liver cell type-biomarker analysis results. Left panel shows heat map of the -log10 corrected P-value for pairs of metabolic biomarker and each of the 10 cell types in the liver. Pairs with FDR < 5% (i.e., *) are colored blue according to their -log_10_ (FDR-corrected *P* value). Non-significant pairs are colored white. Right panel shows bar plot of the number of significant metabolic biomarkers (FDR < 5%) for each cell type in the liver. Abbreviations for metabolic biomarkers are described in Supplementary Table 1. Hep: hepatocytes, HSC: hepatic stellate cells, Mac: Macrophage, and LESC: Liver sinusoidal endothelial cells.

To pinpoint specific liver cell-types linked to the 222 liver-associated metabolic biomarkers, we integrated the GWAS results from BBJ1-180K and the European-specific meta-analysis results from UKB-EBB with publicly available single-nucleus RNA sequencing (snRNA-seq) data from healthy human liver^28^. This snRNA-seq analysis identified 10 cell clusters based on gene expression patterns. Applying MAGMA to perform gene-level association tests between the 222 metabolic biomarkers and 10 clusters, we identified 350 pairs (197 metabolic biomarkers and 10 clusters) (FDR < 5%) (Fig. 3b and Supplementary Table 13). Of the 10 clusters, hepatocytes near the periportal vein showed the largest number of significant associations (151 pairs). This was followed by hepatic stellate cells (HSC) near the periportal vein clusters (HSC 1, 58 pairs) and hepatocytes near the central vein (52 pairs). In contrast, interzonal hepatocytes showed only seven pairs, suggesting a preferential involvement of perivascular hepatocytes in the genetic regulation of the circulating analyzed metabolic biomarkers.

### Associations between blood metabolic biomarker levels and disease risk

Leveraging metabolomic and clinical data from BBJ1-180K, we investigated the relationships between 248 metabolic biomarker levels measured at the blood sampling date (baseline) and the incidence of 19 diseases with at least 20 events after blood sampling. Using Cox proportional hazards models adjusted for sex (excluding prostate cancer) and age at blood sampling, we estimated hazard ratios (HRs) for 4,701 pairs (248 metabolic biomarkers and 19 disease onsets). As a result, 566 pairs (216 metabolic biomarkers and 11 diseases) showed significant associations after Bonferroni correction (*P* value < 0.05/(20 × 19); see Methods) (Supplementary Table 14).

To replicate these 566 pairs, we examined the estimated HRs of 122 matched biomarker-disease pairs in UKB^10^. Of these, 64 pairs were also significant in the same direction (*P* value < 0.05/122 in UKB, after Bonferroni correction) (Supplementary Table 15), with a high Pearson correlation coefficient (*r* = 0.95). By disease, we replicated 39 pairs for myocardial infarction, 17 pairs for ischemic stroke, three pairs for rheumatoid arthritis, one pair for lung cancer, and one pair for cataract. Of the 17 replicated metabolic biomarkers for ischemic stroke, 16 (high density (HDL)-related biomarkers, albumin, and histidine) were also replicated for myocardial infarction (Supplementary Fig. 2), suggesting that these metabolic biomarkers play a broad role in cardiovascular diseases in both populations.

Finally, to examine cross-ancestry disease prediction based on baseline metabolic biomarker levels, we performed an IVW meta-analysis of 2,971 matched pairs (248 metabolic biomarkers and 12 diseases) across BBJ1-180K and UKB. This analysis identified 891 significant pairs (237 metabolic biomarkers and 11 diseases) (*P* value < 0.05/1000, as defined in a previous study^10^), including 47 additional associations (42 metabolic biomarkers and 10 diseases) that were not significant in either BBJ1-180K or UKB alone (Supplementary Table 15).

### Prediction of disease onsets using metabolomic scores

To evaluate the predictive ability of multiple metabolic biomarkers for disease risk in BBJ1-180K, we constructed disease-specific metabolomic scores for five diseases (ischemic stroke, myocardial infarction, lung cancer, cirrhosis, and colorectal cancer). These scores were derived by integrating the baseline metabolomic data from BBJ1-180K with the pre-determined weights of 36 metabolite levels for each disease derived from UKB11. We then assessed the utility of these scores over a 10-year follow-up period in BBJ1-180K. A log-rank test revealed significant differences in disease risk between individuals in the top 10% ranked by metabolomic score (the high-risk group) and the remaining 90% for myocardial infarction and cirrhosis (*P* value < 0.05/5, after Bonferroni correction) (Fig. 4a and Supplementary Table 16). Using Cox proportional hazard models adjusted for sex and age at blood sampling as covariates, we estimated HRs for the high-risk group, showing that cirrhosis and myocardial infarction had significant HRs (HR for myocardial infarction = 2.5, HR for cirrhosis = 8.2, *P* value < 0.05/5, Supplementary Fig. 3 and Supplementary Table 17).

**Fig. 4.**
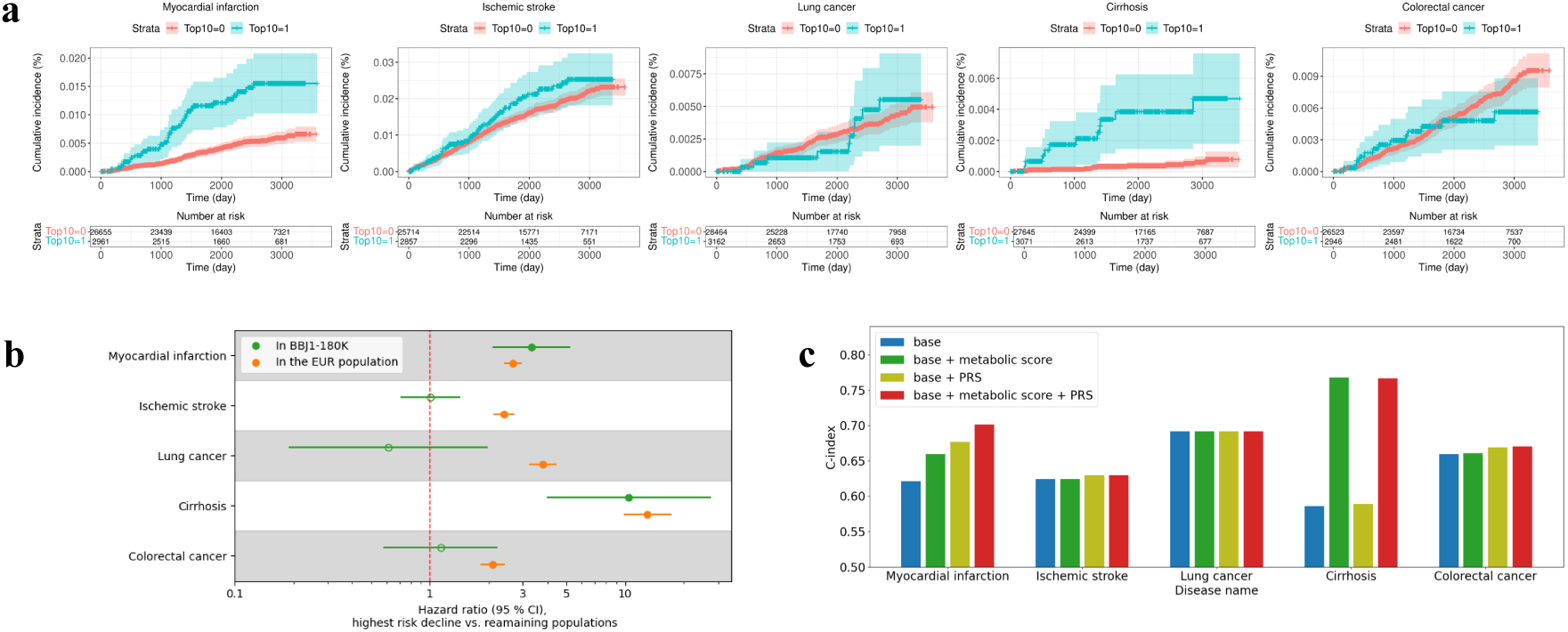
Predictive performance of the metabolomic scores for five diseases. **a**, Kaplan-Meier curves comparing individuals in the top 10% high-risk group with the remaining 90% of the participants in BBJ1-180K. Top10: The top 10% high-risk. **b**, Four-year HRs for the metabolomic scores applied to the top 10% high-risk group for each of the five diseases. Each dot represents the point estimate of HR from Cox proportional hazards model, and the horizontal error bars denote the 95% confidence intervals. Dots are filled when the *P* value is < 0.05/5; otherwise, they are left unfilled. **c**, Comparisons of the C-index for five diseases using three models: base (= age + sex), base + metabolomic score, base + PRS model, and base + metabolomic score + PRS. 10-year follow-up data are used to calculate the C-index.

To compare our findings with those from the predominantly European populations using four-year follow-up data, we repeated the analysis with the same follow-up period. We observed consistent results: the high-risk group had significantly higher risks of myocardial infarction and cirrhosis (*P* value < 0.05/5, after Bonferroni correction) (Supplementary Table 18). When comparing HRs between our study and predominantly European cohorts, we revealed significant heterogeneity for ischemic stroke and lung cancer (Cochran’s Q test *P* value < 0.05/5, after Bonferroni correction) but no heterogeneity for myocardial infarction and cirrhosis (Fig. 4b and Supplementary Table 18).

Next, we compared the predictive performance of metabolomic scores and polygenic risk scores (PRS) for disease risk. PRS were calculated based on GWAS results in BBJ1-180K (samples lacking metabolic biomarker measurement) and evaluated using Cox proportional hazard models adjusted for sex, age, and the top 10 principal components of genetic ancestry. For myocardial infarction, the PRS was significantly associated with disease risk (HR of PRS for myocardial infarction = 3.3, *P* value < 0.05/5, after Bonferroni correction) (Supplementary Table 19) and showed a higher HR than the metabolomic score. We further compared two models: model 1, which incorporated covariates and either the metabolomic score or PRS, and model 2, which incorporated covariates plus both the metabolomic score and the PRS simultaneously. The HRs estimated from model 2 were similar to those from model 1 (Supplementary Fig. 4 and Supplementary Table 20). Metabolomic scores showed significant HRs for cirrhosis and myocardial infarction, while the PRS showed a significant HR only for myocardial infarction. To evaluate the predictive ability, we calculated the concordance index (C-index) for four models: (i) base (age and sex only), (ii) base + metabolomic score, (iii) base + PRS, and (iv) base + metabolomic score + PRS. For cirrhosis and lung cancer, the metabolomic score alone demonstrated the highest predictive value. For myocardial infarction, ischemic stroke and colorectal cancer, the model that combined both metabolomic score and PRS had the highest C-index (Fig. 4c and Supplementary Table 21).

## Discussion

In this large-scale GWAS of 248 blood metabolic biomarkers, we identified eight novel associations in the Japanese population and 28 additional associations in the largest cross-ancestry meta-analysis conducted to date. Statistical fine-mapping prioritized 47 putative causal variants with potential functional consequences, predicted to either alter protein function (n = 24) or regulate gene expression via enhancer elements (n = 23). Integrative gene-level analyses further underscored perivascular hepatocytes as the most genetically relevant cell type contributing to inter-individual variation in circulating metabolic biomarker levels. Moreover, integrating metabolomic and clinical data from BBJ1-180K and comparing our findings with those from primarily European populations showed that metabolomic scores have predictive potential for myocardial infarction and cirrhosis in the Japanese population. Collectively, our study uncovers key genetic variants and cell types that shape inter-individual variation in circulating metabolite levels and provides an atlas linking metabolic biomarkers to disease risk in the Japanese population.

We identified 32 metabolic biomarkers, such as docosahexaenoic acid, ApoA1, phospholipids in small VLDL, cholesteryl esters in small HDL, and LDL size, that were associated with the *CD36* region. Of these, 31 were reported in previous studies, and the association with LDL size represented one of the novel associations identified by this study and was replicated in an independent cohort. Fine-mapping and functional annotation pinpointed rs75326924 in *CD36*, known to cause CD36 deficiency, as the putative causal variant for 15 metabolic biomarkers, including LDL size. CD36 expressed on macrophages recognizes oxidized LDL, a modified form of LDL that tends to form smaller particles^29^. A previous study^30^ reported that macrophages derived from CD36-deficient patients exhibited approximately a 50% reduced oxidized LDL uptake compared to those from healthy individuals. Our results demonstrated that rs75326924 was significantly associated with reduced LDL particle size, potentially attributable to impaired uptake of oxidized LDL due to CD36 deficiency.

In line with previous studies^31,32^, our gene-level analysis showed that the liver is the tissue most genetically associated with metabolic biomarkers, with associations observed for 222 out of 248 biomarkers assessed in this study. To further dissect this relationship, we focused on liver cell types and found that hepatocytes near the periportal and central veins showed significantly greater enrichment than those in the interzonal region. The lack of enrichment in interzonal hepatocytes represents a novel observation. Supporting our findings, a previous study^33^ showed that single-cell disease relevance scores for nine laboratory values, including total LDL and HDL cholesterol, were positively associated with the scores for hepatocytes in both periportal and pericentral regions, whereas our analysis targeted a broader range of metabolic biomarkers and provided a more detailed characterization of lipoproteins in terms of their particle size and components. In adult patients, metabolic dysfunction-associated steatotic liver disease (MASLD), formerly known as non-alcoholic fatty liver disease, has been reported frequently as a liver disease that involves zonation^34^. When comparing metabolic biomarkers associated with the hepatocytes in the periportal and pericentral regions, 24 metabolic biomarkers, including three fatty acids (ratio of omega-3 fatty acids to total fatty acids, ratio of omega-6 fatty acids to omega-3 fatty acids, and ratio of saturated fatty acids to total fatty acids), and two amino acids (glycine and glutamine), showed significant genetic associations exclusively in the pericentral region. Fatty acids are well-known risk factors of MASLD^35^, and glycine and glutamine, key substrates for glutathione synthesis, were also reported to be associated with MASLD development^36^. Together, our findings reveal that the genetic architecture of blood metabolic biomarkers is more tightly linked to perivascular hepatocytes than interzonal ones and provide evidence that some metabolic biomarkers, known to be related to the development of MASLD, are genetically associated with the hepatocytes in the pericentral region.

By integrating metabolomic profiles with longitudinal clinical information from BBJ1-180K, we identified 566 biomarker-disease onset pairs with predictive potential. Of the 122 testable pairs, 64 were successfully replicated in UKB. Inverse associations between cardiovascular disease onset and HDL cholesterol or albumin have been well documented in previous studies^37–39^ and were replicated in the present study. While the protective role of histidine in cardiovascular disease onset has previously been reported only in the European or African populations^10,40,41^ to our knowledge, its association with reduced risk of myocardial infarction was consistently observed in both the BBJ1-180K and UKB studies. Histidine is an essential amino acid with antioxidant and antiinflammatory properties^42^, and its pivotal physiological functions may contribute to a cardioprotective effect across diverse ancestral populations.

We evaluated the predictive ability of metabolomic scores for five disease risks in BBJ1-180K using pre-determined weights derived from UKB metabolomic data. Our results showed significant HRs for cirrhosis and myocardial infarction, suggesting that the prediction model developed in the European population remains effective in the Japanese population. Notably, the metabolomic score outperformed the PRS in predicting the risk of cirrhosis. In contrast, we observed substantial heterogeneity in HRs for ischemic stroke and lung cancer compared with the corresponding European results, whereas a previous study12 reported no such heterogeneity for these two diseases across three European cohorts. The heterogeneity identified here may reflect differences across ancestral populations, such as the subtype distribution for ischemic stroke and lung cancer^43,44^.

A primary limitation of our study is the relatively small number of metabolic biomarkers analyzed. Although NMR spectroscopy is relatively cost-effective and provides consistent quantification, it captures fewer traits than MS, potentially underrepresenting the broader human metabolome, particularly lipid species. Furthermore, among 14,266 95% CSs identified by cross-ancestry fine-mapping, we found that 3099 CSs (21.7%) did not contain a variant with PIP > 0.5 of being ancestry-specific or shared. This likely reflects the smaller sample size in BBJ-180K (N = 37,969) compared to that in the UKB-EBB Europeans (N = 599,249). Therefore, although this study, the largest metabolomic GWAS in East Asian populations to date, contributes to findings across ancestral populations, further expansion of sample sizes in non-European populations is required to more finely understand the genetic architecture of metabolic biomarkers.

In summary, by integrating metabolomic, genomic, and clinical data from BBJ1-180K and comparing our findings with those from primarily the European population, we have elucidated the genetic architecture of metabolic biomarkers and demonstrated their predictive potential for disease onset across ancestral populations.

## Methods

### Study participants

All samples used in this study were collected from BBJ, a multicenter, hospital-based cohort in Japan. From fiscal year 2003 to 2007, BBJ collected DNA, serum, and clinical information from approximately 200,000 patients in 66 hospitals across 12 medical institutions as the first cohort (BBJ1). We obtained informed consent from all participants by following protocols approved by their institutional ethical committees. We obtained approval from the ethics committee of the Institute of Medical Sciences, The University of Tokyo (approval number 2023-77-0118). We have complied with all of the relevant ethical regulations. Serum metabolic biomarkers were quantified in a total of 45,270 samples from BBJ1. BBJ1 comprised two studies using different genotyping arrays, BBJ1-180K (approximately 180,000 individuals) and BBJ1-12K (approximately 12,000 individuals), as described in our previous paper^45^. No individuals overlapped between the two sub-cohorts (see Genotyping and genomic imputation). All study participants were diagnosed by physicians at the cooperating hospitals with at least one of the 47 target diseases^46^. Among these diseases was amyotrophic lateral sclerosis (ALS); however, none of the subjects included in the current study were diagnosed with ALS (Supplementary Table 22). Clinical information was collected annually through interviews and medical records up to fiscal year 2013.^46^

### Metabolic biomarkers quantification, QC, and preprocessing

Blood samples were collected at the collaborating hospitals of BBJ and stored at -80 °C. Samples were transferred to BBJ in a cooling container with dry ice and stored at -150 °C in BBJ. Samples were thawed at 4 °C for aliquoting into 100 μL and then frozen at -80 °C in BBJ before analysis. The serum samples were shipped to Nightingale Health laboratories in Tokyo in 96-well plates on dry ice in batches of 10,000–20,000 samples. Samples were stored in a freezer at -80 °C at Nightingale Health laboratories after arrival from BBJ laboratory. Before preparation, frozen samples were slowly thawed at +4 °C overnight, and then mixed gently and centrifuged (3 min, 3400 × g, +4 °C) to remove possible precipitate. Aliquots of each sample were transferred into 3-mm outer-diameter NMR tubes and mixed in 1:1 ratio with a phosphate buffer (75 mM Na2HPO4 in 80%/20% H2O/D2O, pH 7.4, including also 0.08% sodium 3-(trimethylsilyl) propionate-2,2,3,3-d4 and 0.04% sodium azide) automatically with an automated liquid handler (PerkinElmer Janus Automated Workstation)^10^. The samples were measured using a 500 MHz NMR spectrometer (Bruker AVANCE IIIHD) by Nightingale Health Japan. Nightingale Health’s proprietary software (quantification library 2020) was used to quantify 250 serum metabolic biomarker levels of 45,270 samples from BBJ1 (Supplementary Table 1). These metabolic biomarkers were classified into 108 directly measured metabolites (non-derived biomarkers) and 142 biomarkers, sum and/or ratio of the non-derived biomarkers (derived biomarkers), following the protocol described in a previous study^47^ (Supplementary Table 1).

We performed QC on 45,147 samples from BBJ1-180K (N = 40,659) and BBJ1-12K (N = 4,488). First, we excluded glycerol and 3-hydroxybutyrate, which showed missing rates exceeding 90% across BBJ1-180K and BBJ1-12K, leaving 248 biomarkers for the present study.^47^ We excluded samples if they lacked age information, were under 18 years old, lacked imputation data (due to failure in genotype QC or overlap with the imputation reference panel), or had a diagnosis date for any of the initial 47 target diseases more than six months after the blood sampling date. Samples with over 90% missing biomarker values were also excluded. If individuals had multiple serum measurements, only the sample corresponding to the earliest blood sampling date was retained.

Following these basic QC procedures applied across all analyses, we additionally applied analysis-specific QC steps tailored for metabolomic GWAS and for the survival analysis as described below.

Metabolomic GWAS was conducted after accounting for potential confounding by disease status or medication use. We reviewed clinical information recorded within six months before or after blood sampling for conditions or medications listed in the exclusion criteria (Supplementary Table 23).

Following a previously proposed QC protocol^47^, we first estimated and adjusted the impact of technical variability (metabolomic measurement batches and time from loading to measurement) during the quantification of non-derived biomarkers in each study (N_BBJ1-180K_ = 40,764; N_BBJ1-12K_ = 4,495). We then recalculated derived biomarkers using the R package ukbnmr (v.2.2.1). Metabolic biomarker values beyond ± 4 standard deviations from the mean were set to missing. These steps resulted in 37,969 samples for BBJ1-180K and 3,528 samples for BBJ1-12K.

For survival analysis between metabolic biomarker(s) and disease risk, we excluded samples whose initial date of interviews or medical records was more than six months after the blood sampling date, or who were diagnosed with the target or related diseases (Supplementary Table 24) before or within six months after blood sampling. For the analysis of prostate cancer, we used a subset comprising only men. When building metabolomic scores for disease prediction, we excluded samples with any missing values among the 36 metabolic biomarkers and used the remaining samples to calculate metabolomic scores. These biomarkers have been clinically validated and certified for diagnostic use in the NMR metabolomics assay to facilitate rapid clinical translation^10^. Following a previous study^11^, we estimated a physiological glucose concentration based on glucose and lactate (adjusted glucose concentration = glucose concentration + 0.38 × lactate concentration) to account for potential glucose degradation during sample preparation. Afterward, metabolic biomarker values exceeding ± 4 SDs from the mean were treated as missing. Metabolic biomarker levels were subsequently log1P-transformed and standardized to Z-scores (mean = 0 and variance = 1).

### Genotyping and genomic imputation

All BBJ1-180K samples were genotyped using Illumina HumanOmniExpressExome BeadChip or a combination of Illumina HumanOmniExpress and HumanExome BeadChips. BBJ1-12K samples were genotyped using Infinium Asian Screening Array BeadChip. QC for the genome data of samples in each cohort followed the protocol described in a previous study^48^. For sample-level QC, individuals with a call rate of less than 98% were removed. After removing duplicates and twins within each cohort based on estimated identical-by-descent (IBD) sharing and retained only the samples with the highest call rate, we re-estimated IBD sharing to confirm no sample overlapping between the two sub-cohorts (BBJ1-180K and BBJ1-12K). For genotype QC, we excluded variants with a genotype call rate < 99%, heterozygous counts < 5, Hardy-Weinberg equilibrium *P* value < 1.0 × 10^−6^, or for BBJ1-180K alone, a concordance rate < 99.5% or a non-reference discordance rate ≥ 0.5% between array genotypes and whole-genome sequence data in the same individuals (N = 939). Pre-phasing was performed using Eagle (v2.4.1)^49^. Imputation was conducted using a reference panel constructed from whole-genome sequence data of Japanese ancestry (n = 3,256) combined with the 1KGP3 version 5 (n = 2,504)^50^. All genomic coordinates are in hg19 (GRCh37), except for the VEP downstream analysis. The imputation was carried out using Minimac4 (v1.0.2; https://github.com/statgen/Minimac4). Variants with an imputation quality *R*^2^ < 0.7 or MAF < 0.1% were excluded.

### Metabolomic GWAS using samples from BBJ1-180K

After applying an inverse rank normal transformation to the post-QC metabolic biomarker levels, we performed a GWAS using the linear mixed model implemented in BOLT-LMM (v2.4.1)^15^. The model included age at blood sampling, sex, the top 10 principal components of genetic ancestry, and the presence or absence of 46 registered diseases developed before or within six months after blood sampling as covariates. For the discovery GWAS, we used samples from BBJ1-180K (N=37,969). We calculated minus log_10_ (*P* value) based on *χ*^2^.

### Principal component analysis of metabolic biomarker levels

Using the Python package scikit-learn (v.0.24.2), we performed principal component analysis (PCA) on the inverse rank-normalized levels of 248 metabolic biomarkers in 9,444 samples from the QC-processed BBJ1-180K that had no missing values. Default parameters were used for the analysis. The top 20 principal components cumulatively explained 95% of the variance in the 248 metabolic biomarkers. Considering that this study was conducted on multiple correlated metabolic biomarkers, we adjusted the conventional genome-wide significance level (5×10^−8^) by these 20 principal components, yielding a study-wide corrected significance level^51^.

### Functional annotation of variants by VEP

To take advantage of more up-to-date annotations, we lifted over the coordinates of the GWAS variants from hg19 to hg38 using the Python package gwaslab (v.3.5.1; https://github.com/Cloufield/gwaslab)^52^ and then annotated the variants with VEP (https://useast.ensembl.org/info/docs/tools/vep/index.html)^53^ version 99, using GENCODE v49 on hg38. We adopted the Ensembl severity ranking (https://asia.ensembl.org/info/genome/variation/prediction/predicted_data.html)^31^ and defined functional variants as those containing any of the following terms: splice_acceptor_variant, splice_donor_variant, stop_gained, frameshift_variant, stop_lost, start_lost, transcript_amplification, inframe_insertion, inframe_deletion, missense_variant, protein_altering_variant, splice_region_variant.

### Definition of associated regions and novel associations

We identified variants with *P* value below the study-wide corrected significance level for each metabolic biomarker and defined 1 Mb windows centered on these variants as associated loci. To capture all associated regions across metabolic biomarkers, we merged partially overlapping windows to eliminate redundancy. Within each merged region, the variant showing the strongest association with a given metabolic biomarker was designated the lead variant. Among these lead variants, the one with the smallest *P* value across all associated biomarkers was selected as the cross-metabolic biomarkers’ lead variant for the region (Supplementary Fig. 4).

To name each region, we first examined whether the cross-metabolic biomarker lead variant and variants in LD with it (r^2^ > 0.6) were annotated as functional variants. We assessed LD in East Asians from 1KGP3 for BBJ1-180K GWAS results and LD in both East Asians and Europeans from 1KGP3 for cross-ancestry meta-analysis results. If functional variants were identified, we adopted the gene(s) affected by these variants as the region name. When multiple functional variants were available, we selected the gene associa the variant that had the highest Combined Annotation-Dependent Depletion score (v.1.6)^54^. If no functional variant was identified, we used the name(s) of the closest gene(s) of the lead variant. A lead variant for a metabolic biomarker was considered a novel association when it was located more than 1 Mb away from any previously reported associated variants (*P* value < 5×10^−8^ or stronger study-specific thresholds). We consulted the GWAS catalog (v.2024-12-19)^55^ and additional studies not listed in the GWAS catalog (six to 200 studies for each GWAS; Supplementary Table 25), including metabolomic GWAS results from Europeans^7,56–58^ and East Asians^6,7,59–61^. The experimental factor ontology (EFO) IDs were used to comprehensively map the same or similar phenotypes in the GWAS catalog.

### Replication for novel associations

In BBJ-12K, we tested variant-metabolic biomarker associations that were identified as novel in either the GWAS in BBJ1-180K or the cross-ancestry meta-analysis, using a linear model implemented in PLINK2 (v2.00a3LM; https://www.cog-genomics.org/plink/2.0/)^62^ after excluding related individuals (N = 3,522). To evaluate these novel associations, we performed a sign test using replication results from BBJ-12K for associations identified in BBJ1-180K, and using both BBJ-12K and an additional previous large-scale cross-ancestry meta-analysis^3^ for those identified in the present cross-ancestry meta-analysis. This test evaluated whether the proportion of variants with matching effect directions exceeded what would be expected by chance. We performed a binomial test to assess whether the observed number of concordant effect directions in both studies was significantly greater than 50%. A one-sided test *P* value < 0.05 was considered evidence of replication.

### Stepwise conditional analysis

To identify independent signals within and around the associated region (± 500 kb), we performed a stepwise conditional analysis. In each iteration, we regressed the metabolic biomarker on every variant while adjusting for the most significant variant in the region (the index variant) and for the covariates used in the original GWAS. The procedure was repeated until no variant reached the genome-wide significance threshold (*P* value < 5×10^−8^). This analysis identified 1,443 additional associations (213 index variants with *P* value < 5 × 10^−8^ and 236 metabolic biomarkers) (Supplementary Table 26).

### Fine-mapping in BBJ1-180K

For each of the 3,583 independent signals identified by GWAS and stepwise conditional analysis, excluding the MHC region (chromosome 6: 25-36 Mb), we defined a 2-Mb genomic region centered on the lead variants or index variants. Overlapping regions for the same metabolic biomarker were merged, resulting in 2,111 distinct regions ranging from 2.0 Mb to 3.9 Mb. To pinpoint causal variants, we performed statistical fine-mapping using SuSiE-RSS (v.0.12.35)^16^, allowing up to 10 CSs per region. For genetic relationship inference, we used the KING framework implemented in PLINK2 to compute the KING kinship coefficient. Individuals with a KING kinship coefficient >0.0884 were excluded, resulting in a set of 37,288 unrelated individuals from the 37,969 used in this study within BBJ1-180K. Using this dataset, we constructed the LD matrix using LD store2 (v.2.0; http://www.christianbenner.com) for the fine-mapping with SuSiE-RSS.

### Nomination of genes for non-coding variants by the ABC model

We used the genome-wide mapping of enhancer-gene connections in 131 human cell or tissue types predicted by the ABC model. The dataset included the element-gene links with ABC scores >= 0.015 (https://mitra.stanford.edu/engreitz/oak/public/Nasser2021/AllPredictions.AvgHiC.ABC0.015.minus150.ForABCPaperV3.txt.gz)^18^. When a putative non-coding causal variants falls within a regulatory element, we assume that it influences enhancer activity and, consequently modulates the expression of the associated target genes.

### Cross-ancestry meta-analysis

Using the metabolomic GWAS results from BBJ1-180K and the meta-analysis results of six ancestral populations from UKB-EBB, we conducted fixed-effects meta-analyses with the IVW method implemented in METAL (v.2011-03-25, https://csg.sph.umich.edu/abecasis/Metal/download/)^22^. Variants were retained only if they were present in both BBJ1-180K and UKB-EBB.

### Comparison of genetic effect sizes across ancestral populations

Using GWAS results from BBJ1-180K, the European-specific meta-analysis from UKB-EBB, as well as the GWAS results from five ancestral populations (the Central/South Asian, African, East Asian, Middle Eastern, and Admixed American population) within the UKB, we investigated the correlations of the genetic effect sizes of extracted lead variants.

We first selected lead variants from BBJ1-180K with MAF > 1% in both BBJ1-180K and each of the six comparison studies from UKB or UKB-EBB, resulting in 1,638 associations for the European population, 1,680 for the Central/South Asian population, 1,660 for the African population, 1,846 for the East Asian population, 1,659 for the Middle Eastern population, and 1,642 for the Admixed American population (Supplementary Table 8).

Next, we defined associated regions across metabolic biomarkers based on the European-specific meta-analysis or GWAS results from each population. We again extracted lead variants with MAF > 1% in both BBJ1-180K and each comparison study, resulting in 32,187 associations for the European population, 749 for the Central/South Asian population, 300 for the African population, 241 for the East Asian population, 18 for the Middle Eastern population, 6 for the Admixed American population (Supplementary Table 9).

We then calculated Pearson correlation coefficients to assess the concordance of genetic effect sizes between BBJ1-180K and each of the six ancestral populations from UKB-EBB or UKB.

### Multi-ancestry fine-mapping

For the 2,111 regions constructed for fine-mapping with SuSiE-RSS, we performed multi-ancestry fine-mapping to identify causal variants across ancestries with MESuSiE (v.1.0)^24^, using GWAS results from the East-Asian specific BBJ1-180K and the European-specific meta-analysis from UKB-EBB, and LD matrices from BBJ1-180K and the unrelated British-ancestry individuals from UKB (N = 337,491)^63^. MESuSiE models genetic effects under both ancestry-shared (non-zero effect in both ancestries) and ancestry-specific conditions and calculates a PIP for each variant in each condition. Following the previous study^24^, we excluded variants with MAF < 0.1% in European-specific meta-analysis and strand-ambiguous variants in both studies. Variants were classified as shared causal variants if PIP > 0.5 in both East Asian and European populations, East Asian-specific causal variants if PIP > 0.5 in the East Asian population only, and European-specific causal variants if PIP >0.5 in the European population only. We allowed up to 10 signals per region.

### Gene-level association analysis between metabolic biomarkers and tissue or cell types

Using the GWAS results from BBJ1-180K and the European-specific meta-analysis from UKB-EBB, we performed gene-based analysis using MAGMA (v.1.08) implemented in FUMA (v.1.6.2) with default parameters. We used the LD reference panel constructed from genomic data of the 1KGP3 EAS or EUR population that matched the target population. We performed MAGMA^26^ gene-property analysis implemented in the SNP2GENE method of FUMA (v.1.6.2)^27^ integrating the gene-based analysis results for 248 metabolic biomarkers in BBJ1-180K and UKB-EBB with the transcriptome data from the GTEx consortium v.8 across 30 tissues^25^. Tissue-metabolic biomarker pairs with a one-sided test *P* value < 0.05/(20×30) in each study were considered significant after Bonferroni correction, where 20 represents the number of metabolic principal components explaining 95% of the variance in the 248 metabolic biomarkers, and 30 is the number of the analyzed tissue types.

To identify cell types in the liver associated with the metabolic biomarkers, we obtained a publicly available snRNA-seq dataset of the human liver^26^. In the snRNA-seq analysis, approximately 90,000 cells from three healthy human livers were analyzed and 10 cell populations (clusters) were identified based on gene expression patterns. The top 10% highly associated genes compared with the other clusters based on fold-change were considered cluster-type-specific gene sets. Subsequently, for the 222 metabolic biomarkers associated with the liver in the tissue enrichment analysis in UKB-EBB, we performed gene-set enrichment analysis using MAGMA with the gene-based analysis results from BBJ1-180K and UKB-EBB together with cluster-specific gene sets. We used 1KGP3 EAS and EUR population datasets as reference panels. To increase statistical power, we performed an IVW meta-analysis by combining statistics from BBJ1-180K and UKB-EBB. Cluster-metabolic biomarker pairs with FDR < 5% were considered significant.

### Disease risk prediction by each metabolic biomarker

We examined the associations between 248 metabolic biomarkers and 19 disease incidences, each with at least 20 events, based on 34,933 samples from BBJ1-180K. Disease onset was defined as the first recorded diagnosis date after blood sampling. We estimated HRs for the association between Z-normalized metabolic biomarkers and disease incidence using Cox proportional hazards model, adjusting for age at blood sampling and sex (except for prostate cancer, where only male samples were analyzed). Disease-metabolic biomarker pairs with a *P* value < 0.05/(20×19) were considered significant, where 20 is the number of metabolic principal components explaining 95% of the variance in the 248 metabolic biomarkers and 19 is the number of analyzed diseases.

For comparisons and meta-analysis with results from the UKB results^10^, we used the publicly available summary statistics (https://nightingalehealth.com/atlas), which estimated HRs using Cox proportional hazards models adjusted for age at blood sampling, sex and UKB assessment centre. Diseases in BBJ and UKB were matched based on 10th version of International Classification of Diseases (ICD-10) codes (Supplementary Table 27). When a disease-biomarker pair in BBJ corresponded to multiple pairs in UKB, we retained the one with the smallest *P* value in UKB. We performed an IVW meta-analysis to combine results across cohorts.

### Disease risk prediction by metabolomic score

Among the 12 previously analyzed diseases, we evaluated the predictive ability of metabolomic scores for the onset of five diseases (ischemic stroke, myocardial infarction, lung cancer, cirrhosis, and colorectal cancer) for which diagnosis dates were available in BBJ1-180K. Metabolomic scores were calculated by multiplying the Z-normalized metabolic biomarker levels in BBJ1-180K by the corresponding coefficients derived from UKB^11^.

To obtain GWAS results for constructing the PRS models, we performed GWAS for five diseases using individuals without metabolic biomarker measurements in BBJ1-180K (N_total_ = 139,952) with SAIGE (v.1.1.9)^64^ (Supplementary Table 28). For lung cancer and colorectal cancer, we used individuals without any cancers among BBJ target diseases as controls. We included age, age^2^, sex, age × sex, age^2^ × sex, and the top 10 genetic principal components as covariates. The genomic inflation factors *λ*_GC_ indicated no substantial inflation (*λ*_GC_ = 1.02-1.04). We employed PRS-CS to compute PRS models using HapMap3 SNPs with an EAS LD reference panel from the 1KG. Global shrinkage parameters were inferred from the data by PRS-CS using a fully Bayesian approach (PRS-CS-auto).

Individuals were stratified into the top 10% (high-risk group) and the remaining 90% ranked by their metabolomic score or PRS. Kaplan-Meier curves stratified by the score were estimated with the R package survival (v.3.5.7). Using Schoenfeld’s residuals, we did not reject the proportional hazards assumption for any of the five diseases (*P* value > 0.05/5). We also estimated HRs for the high-risk group within four or 10 years after blood sampling using the Cox proportional hazards model, adjusting for sex and age as covariates. Age at blood sampling was also Z-normalized. We compared the HRs estimated in this study with those from meta-analyses of UKB, EBB, and Finnish THL Biobank. In each study, HRs were estimated using Cox proportional hazards models, adjusting for age and sex as covariates using four years of follow-up data. Furthermore, we computed the C-index to evaluate the predictive ability of the following four models: (1) base (age + sex), (2) base + metabolomic score, (3) base + PRS, and (4) base + metabolomic score + PRS.

## Data availability

Summary statistics for GWAS in BBJ1-180K and the cross-ancestry meta-analysis of 248 metabolic biomarkers are available in the GWAS catalog (accessions GCST90709281 - GCST90709776, Supplementary Table 29). Summary statistics for ancestry-specific GWAS in UKB or UKB-EBB and the cross-ancestry meta-analysis across these studies are available in the GWAS catalog. Genotype and metabolic biomarker data in BBJ1-180K and BBJ1-12K were deposited at the NBDC Human Database (Genotype data in BBJ1-180K, research ID: hum0014 JGAS000114; Genotype data in BBJ1-12K and metabolic biomarker data, research ID: hum0311 JGAS000412 and JGAS000561). The imputation reference panel constructed using whole genome data of 3,256 Japanese individuals from BBJ and 2,504 individuals from 1KG was also deposited at the NBDC Human Database (research ID: hum0014 JGAS000746).

## Code availability

In the present study, we used publicly available software and packages for bioinformatics analysis. The details were described in Methods.

## Acknowledgments

We thank Peter Würtz and Machiko Usui (Nightingale Health) for their valuable support, and we acknowledge Nightingale Health Japan Plc for providing the metabolic biomarker data through a collaboration with BBJ. We also thank Chikashi Terao, Kohei Tomizuka, and Satoshi Koyama for their generous support. We are also gratetful to all the participants and staff of the BBJ project. This study was supported by the Ministry of Education, Culture, Sports, Science, and Technology (MEXT) of the Japanese government and the Japan Agency for Medical Research and Development (AMED) under grant Numbers JP18km0605001 / JP23tm0624002 (the BioBank Japan project), JP19km0405215, and JP223fa627011, and JST SPRING GX, Grant Number JPMJSP2108. The super-computing resource was provided by Human Genome Center, the Institute of Medical Science, the University of Tokyo (http://sc.hgc.jp/shirokane.html).

## Ethics declarations

Competing interests

M.K. is a consultant for Takeda Pharmaceutical Co., Ltd. S.N. is an employee of Daiichi Sankyo Co., Ltd. This work was neither part of a collaboration with nor funded by Daiichi Sankyo. Y.K. holds stock in StaGen Co., Ltd. The other authors declare no competing interests.

