## Supplementary Figures 1-5 for "Genetic Architecture of Circulating Metabolic Biomarkers Across Ancestral Populations"

Supplementary materials

Supplementary Figures

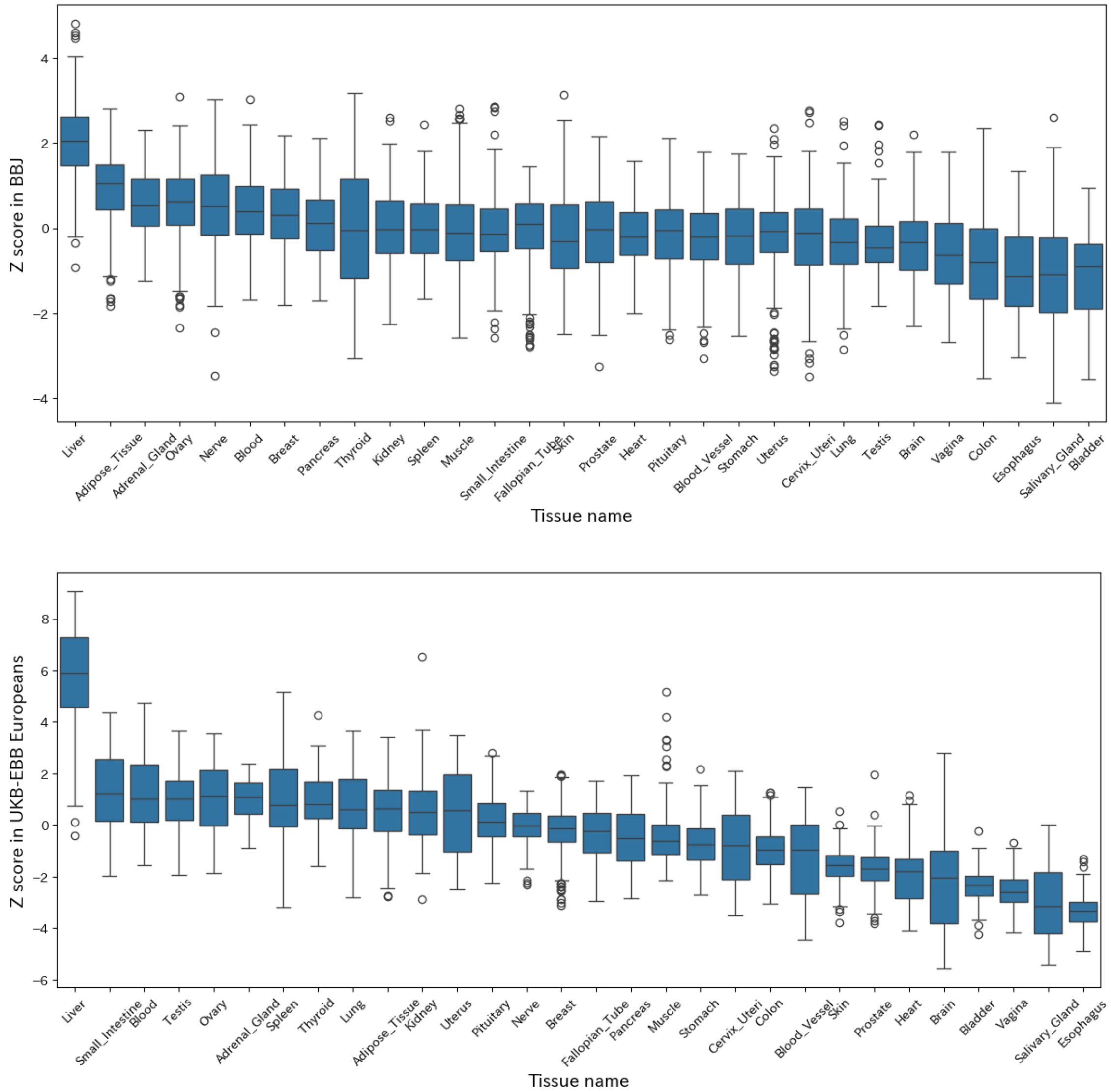


**Supplementary Figure 1 Z scores in gene-level association analysis between 248 metabolic biomarkers and 30 tissues.**

Z scores are estimated using the GWAS results in BBJ (a) and the European-specific meta-analysis results (b). These Z scores are presented as box plots. In each box, the center line indicates the median, and the lower and upper edges correspond to the 25th and 75th percentiles, respectively. Whiskers extend to the smallest and largest values within 1.5 times the interquartile range from the lower and upper quartiles, respectively. Data points outside this range are shown. Tissues along the x-axis are ordered by descending median value.

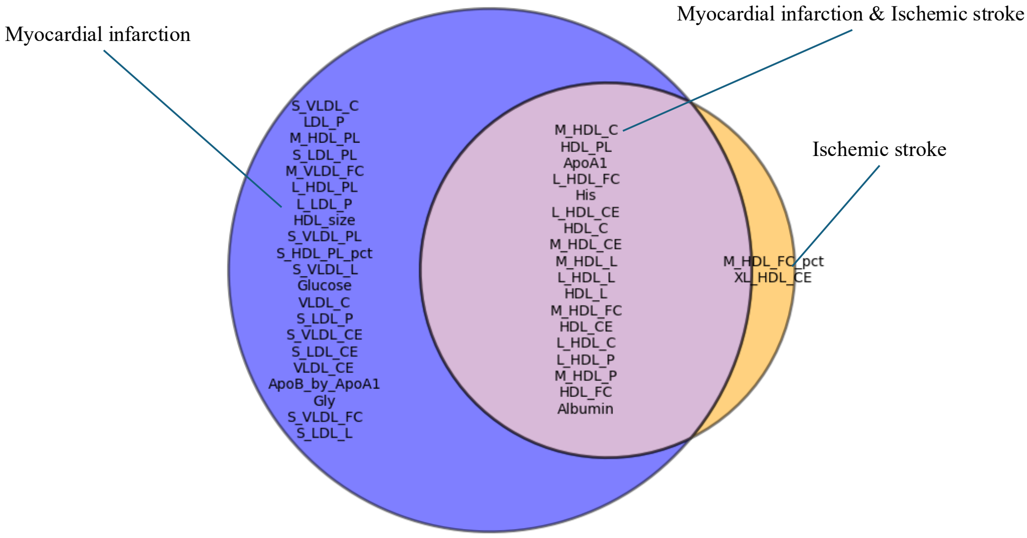


**Supplementary Figure 2 Venn diagram showing the replicated pairs for myocardial infarction and ischemic stroke in UKB.**

Among the pairs identified in BBJ1-180K involving metabolic biomarkers and myocardial or cerebral infarction, the pairs that were replicated in UKB are shown. Abbreviations of metabolic biomarkers are described in Supplementary Table 1.

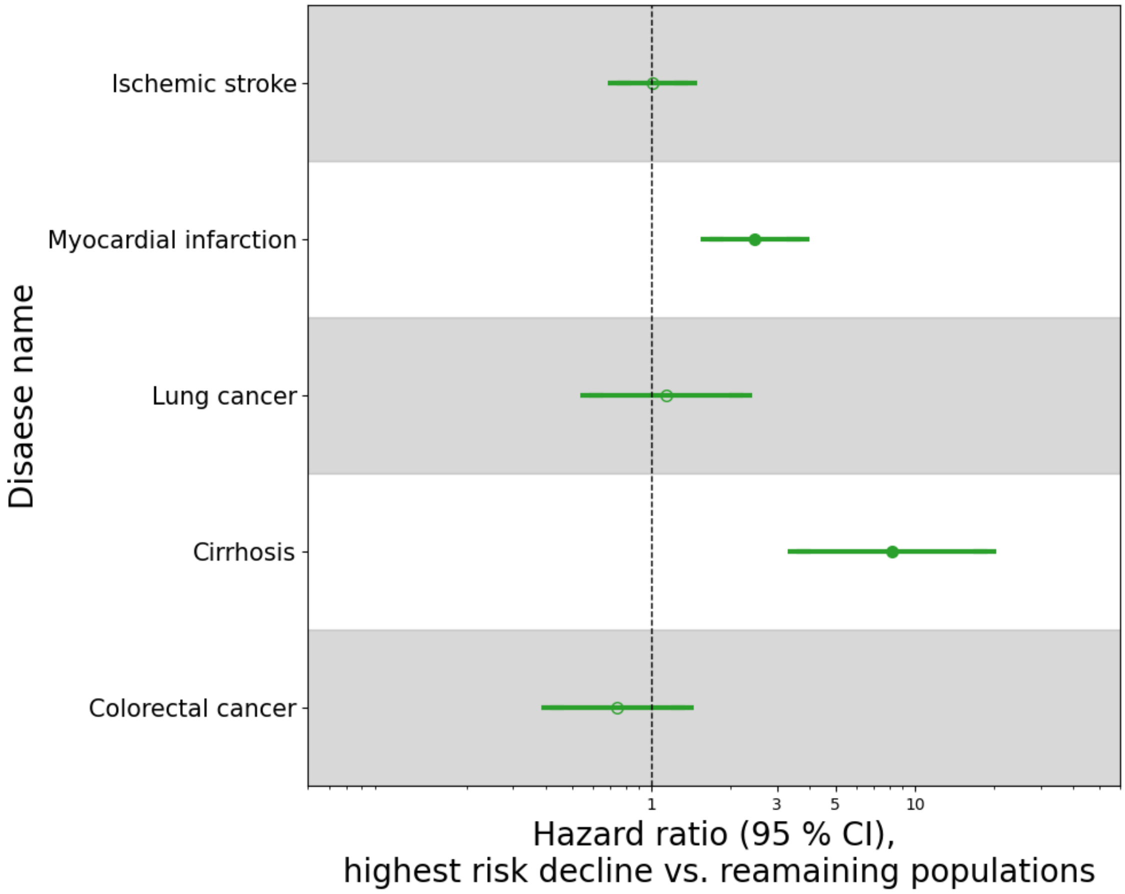


**Supplementary Figure 3 10-year hazard ratios of metabolomic scores for this study's top 10% high-risk group across five diseases.**

Dots represent point estimates of HRs from Cox proportional hazards models, and the horizontal error bars denote the 95% confidence intervals. Dots are filled in if the *P* value was < 0.05/5; otherwise, they are left unfilled.

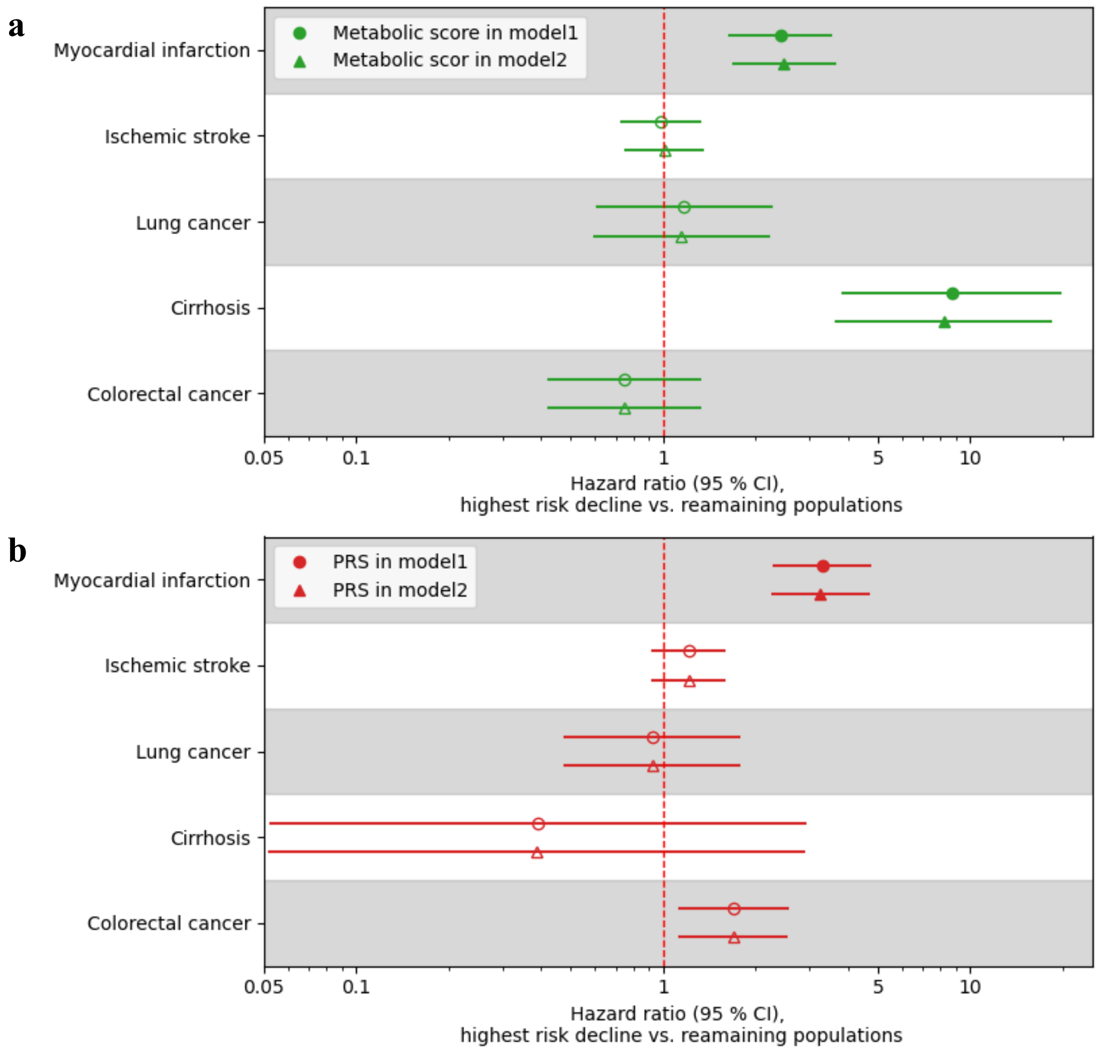


**Supplementary Figure 4 Comparison of HRs of metabolic score and PRS in two models**

10-year HRs of metabolomic scores for the top 10% high-risk group in this study across five diseases in two models (model1: age + sex + metabolic score or PRS + genetic PCA1-10 and model2: age + sex + metabolic score + PRS + genetic PCA1-10). Dots represent point estimates of HRs from Cox proportional hazards models, and the horizontal error bars denote the 95% confidence intervals. Dots are filled in if the *P* value was < 0.05/5; otherwise, they are left unfilled.

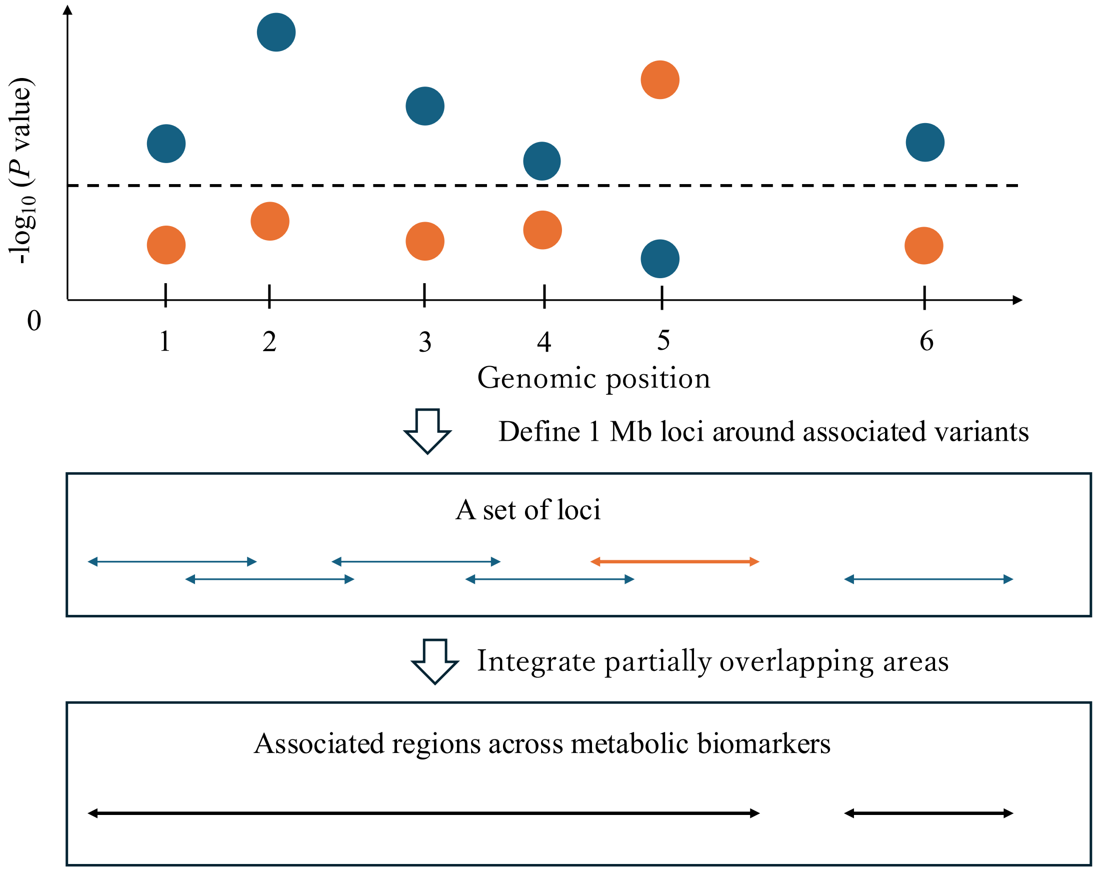


**Supplementary Figure 5 Example of the definition of cross-metabolic biomarker-associated regions**

The top panel displays *P* values from GWAS plotted against genomic positions for metabolite A (blue) and metabolite B (orange). Dashed line indicates a study-wide significance threshold (= $-\log_{10}(5 \times 10^{-8}/20)$). Loci are defined using associated variants with *P* values < a study-wide significance threshold. In this example, loci are defined using variants at genome positions 1, 2, 3, 4, and 6 for metabolite A, and variants at genome position 5 for metabolite B. Consequently, associated regions are defined by merging overlapping regions across metabolites. Ultimately, three associations (three metabolite-specific lead variants, two metabolites) and two regions were counted in this example.
